# Sensitive on-site detection of SARS-CoV-2 by ID NOW COVID-19

**DOI:** 10.1101/2021.04.18.21255688

**Authors:** Eva Krause, Andreas Puyskens, Daniel Bourquain, Annika Brinkmann, Barbara Biere, Lars Schaade, Janine Michel, Andreas Nitsche

**Author notes:** Corresponding author Prof. Dr. Andreas Nitsche, Robert Koch Institute, Centre for Biological Threats and Special Pathogens – Highly, Pathogenic Viruses, Seestraße 10, 13353 Berlin, Germany. These authors contributed equally to this work (Eva Krause, Andreas Puyskens).

## Abstract

Point of care detection of SARS-CoV-2 is one pillar in a containment strategy and important to break infection chains. Here we report the sensitive, specific and robust detection of SARS-CoV-2 and respective variants of concern by the ID NOW COVID-19 device.

## Introduction

The reliable detection of SARS-CoV-2 genomes in clinical specimens is one the most crucial task in the identification of SARS-CoV-2-infected patients, which is a precondition of conducting proper patient management and breaking infection chains [1]. The gold standard of virus diagnostics is the real-time PCR that is highly sensitive, specific and quick and can give semi-quantitative results under appropriate conditions [2][3]. For the detection of SARS-CoV-2 many in-house real-time PCR assays have been published in addition to a plethora of ready-to-use commercially available kits [4–9]. Real-time PCR is suitable to be run as high-throughput approach which takes between 3-4 hours for low numbers of specimens including RNA extraction, but can take up to more than a day for high numbers of specimens due to pre-analytical procedural steps. However, a well-appointed laboratory as well as skilled personnel is required for PCR diagnostics.

Recently, so-called rapid diagnostic tests (RDTs) have reached a status of reliability where they can be applied as self-tests even by non-trained personnel [10–12]. Usually these tests are based on lateral flow assays that specifically recognize viral proteins and can be assessed either visually or with a test-specific device [13]. Commonly, these tests take about 15 minutes after sampling but are not well suited for high-throughput applications. Most importantly, when comparing their positivity rates to real-time PCR results, these RDTs have an analytical sensitivity that is significantly lower than that of real-time PCR [10]. Hence, depending on the scenario, the choice of the best diagnostic approach for SARS-CoV-2 has to balance sensitivity, time to result, throughput and simplicity.

The ID-NOW platform utilizes an isothermal nucleic acid amplification reaction for the qualitative detection of SARS-CoV-2 viral nucleic acids, which suggests a potentially higher sensitivity compared to lateral flow-based RDTs [14]. The time to result is approximately 13 minutes for negative specimens and even less for positive specimens, which is considerably shorter than that of a real-time PCR. The throughput is relatively low as each device can only process one sample at a time. However, there are settings in which a short time to result combined with a relatively high analytical sensitivity are preferred to a high sample throughput. To evaluate whether the ID NOW COVID-19 test can address these demands, we compared the ID NOW COVID-19 test with a standard real-time PCR assay.

## Material & Methods

For the evaluation of the ID NOW COVID-19 test, specimens from primary diagnostics were used in anonymized form. The study obtained ethical approval by the Berliner Ärztekammer (Eth 20/40). These specimens comprised dry or wet nasopharyngeal and/or oropharyngeal swabs sent to the Robert Koch Institute for PCR diagnostics. Wet swabs were kept in transport media, dry swabs were transferred to 1000 µL of PBS. All swabs were vortexed and spun down prior to extraction of 140 µl of the specimen, using the QIAamp Viral RNA Mini Kit and QIAcube Connects with the manual lysis protocol. PCR diagnostics was performed as described previously (under review). Remaining specimen volume was stored at -40°C to - 80°C. In total, 179 SARS-CoV-2-positive specimens were subjected to the ID NOW COVID-19 test. These specimens covered a range of viral loads from 2.3×10^2^ to 7.4×10^8^ SARS-CoV-2 genome copies/mL.

For specificity testing, 36 clinical specimens negative for SARS-CoV-2, but sampled successfully as demonstrated by the presence of human nucleic acid (c-myc CT values between 26 and 38), were used with a PBS background. Furthermore, 40 swab samples taken from various healthy persons every second day and intermittently positive controls were measured to see whether randomly taken specimens give false-positive results. In addition, 16 respiratory clinical specimens previously sampled from patients with influenza-like illness within the German influenza sentinel [15] containing influenza virus A/H1N1 (2009), B/vic, B/yam, H3N2, RSV, HMPV, AdV, HRV, PIV-1, PIV-2, PIV-3, PIV-4, HKU1, HCoV-229E, -OC43 or -NL63 with CT values between 18 and 27 were tested. In total, 92 specimens were analyzed to evaluate the specificity.

Finally, cell culture supernatants of a reference strain (SARS-CoV-2/Italy-INMI1, kindly provided by Antonino DiCaro via the GHSAG network), the B.1.1.7 variant (Robert Koch Institute [RKI] isolate), the B.1.351 SA variant (kindly provided by Thorsten Wolff, RKI) and the Brazil TY7-503 variant of the P.1 lineage (hCoV-19/Japan/TY7-503/2021, kindly provided by Takaji Wakita, National Institute of Infectious Diseases, Japan, via the GHSAG network) grown in Vero E6 cells were used to test whether the ID NOW COVID-19 test detects these variants with a similar sensitivity as that for the SARS-CoV-2 variants circulating during the first months of the pandemic. RNA load was quantified by real-time PCR and variant identity confirmed by sequencing. In total, 12 dilutions (1:2) covering a range of the expected detection limit from 5.5 x 10^5^ genomes per mL to 300 genomes per mL were generated and measured in the ID NOW COVID-19 test. When three consecutive dilutions showed negative results, the analysis was stopped.

## ID NOW COVID-19 testing

Testing was performed using the ID NOW™ COVID-19 test (REF 190-000; Abbott Diagnostics Scarborough Inc., Scarborough, MA, USA) according to the manufacturer’s instructions with the ID NOW™ Instrument (NAT-024; Abbott). Quality control (QC) testing was performed prior to sample testing using the positive swab provided (REF 190-415, Lot: 123496) and a negative patient swab. For sample testing, either 50 µl or 100 µl of the respective sample were added directly to the elution buffer of the sample receiver and resuspended carefully.

## Results

None of the 92 specimens PCR negative for SARS-CoV-2 showed a positive result when analyzed with the ID NOW COVID-19 test, indicating a high analytical specificity for SARS-CoV-2. One out of 40 swab samples of SARS-CoV-2-negative persons showed an invalid result. Repetition of the sampling gave a negative result in the following ID NOW COVID-19 test. Only one of the 36 clinical specimens with a PBS background resulted in an invalid result, which is likely due to the cloudiness of the specimen (that was not causing problems in the real-time PCR). Clinical specimens containing respiratory viruses as listed above remained negative, resulting in an overall specificity of 100%.

For determination of the detection limit of the ID NOW system, in total, 179 clinical specimens positive in the SARS-CoV-2 real-time PCR, with CT values between 14.6 (7.4×10^8^ genome copies per mL) and 36.4 (2.3×10^2^ genome copies per mL), were subjected to the ID NOW COVID-19 test system. One specimen showed an invalid result. Overall, of the remaining 178 specimens, 142 were positive with the ID NOW and 35 specimens were negative (overall sensitivity 80.2%). Considering that according to various studies a patient is infectious with a genome load of >10^6^ copies per mL [16][17][18], reflected by a CT value of <24.7 in our system (n=56), the ID NOW has a sensitivity of 100% for potentially infectious specimens. Binary logistic regression revealed a 50% detection probability at CT 32.3 (95% CI 31.7 to 33.0) representing 3.7 ×10^3^ (95% CI 5.7×10^3^ to 2.3×10^3^) genome copies per mL, with individual specimens detected positive containing only 800 genome copies per mL (figure 1).

**Figure 1:**
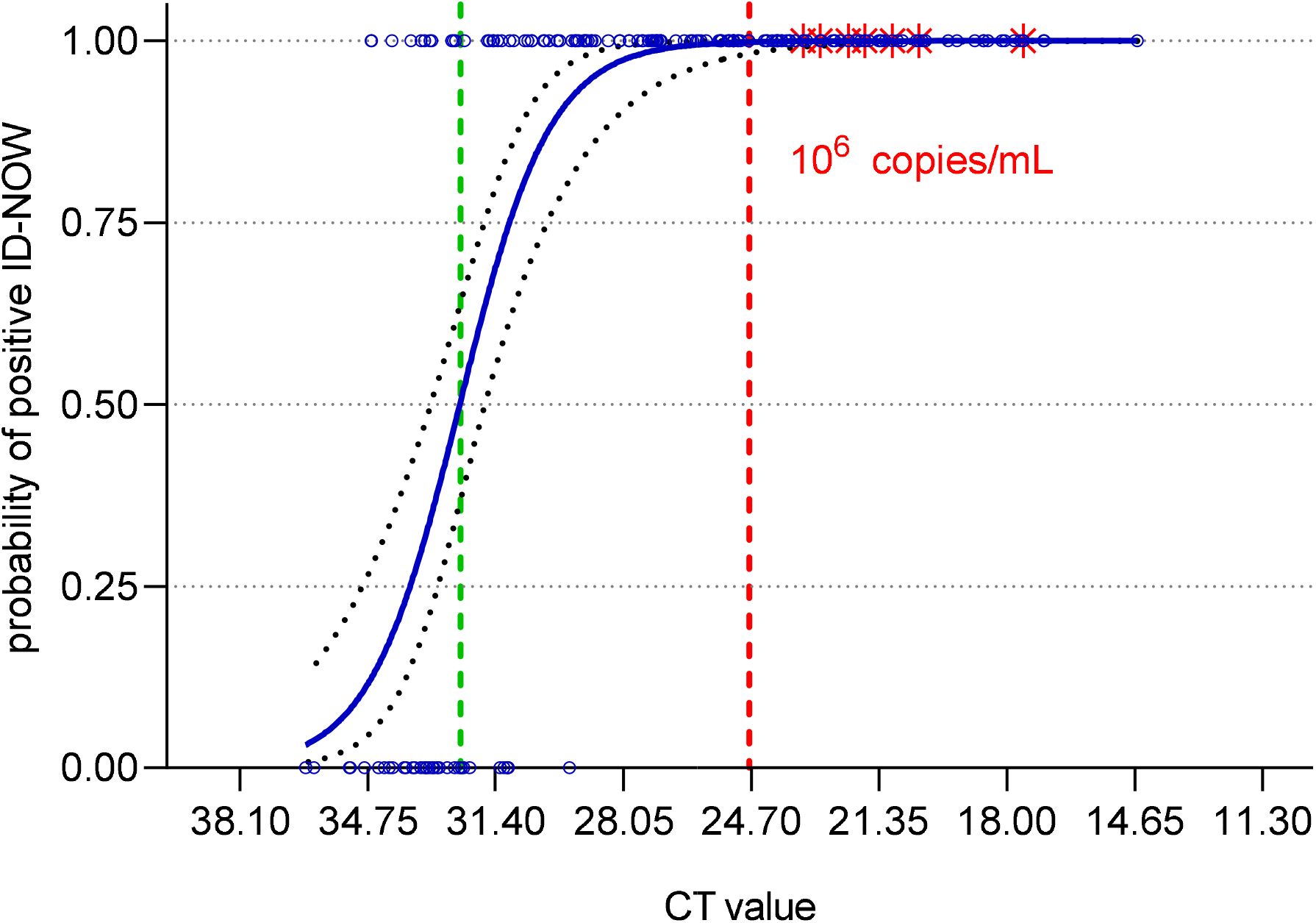
Analytical sensitivity of the ID NOW COVID-19 system as determined by binary logistic regression. Positivity rate as well as the 95% CI are plotted vs the CT value for each specimen (blue circles). Red stars indicate specimens from which SARS-CoV-2 could be grown in cell culture, and the red line shows the virus load of 10^6^ genome copies per mL specimen actually accepted as necessary limit for infectious specimens. The green line indicates the 50% detection probability.

To test whether recently increasingly circulating variants of concern (VOC) can be detected with the ID NOW COVID-19 test, we used cell culture supernatants of the variants listed above. Genome numbers were determined by real-time PCR. Since all analyzed variants show identical sequences in the PCR target regions (E-Gene and orf1ab), cross-comparison of genome numbers by referring to the CT value is appropriate. As shown in figure 2, the detectable genome loads are comparable between the different VOC B.1.1.7, B.1.351 SA, Brz TY7-503 and the reference strain Italy/INMI1.

**Figure 2:**
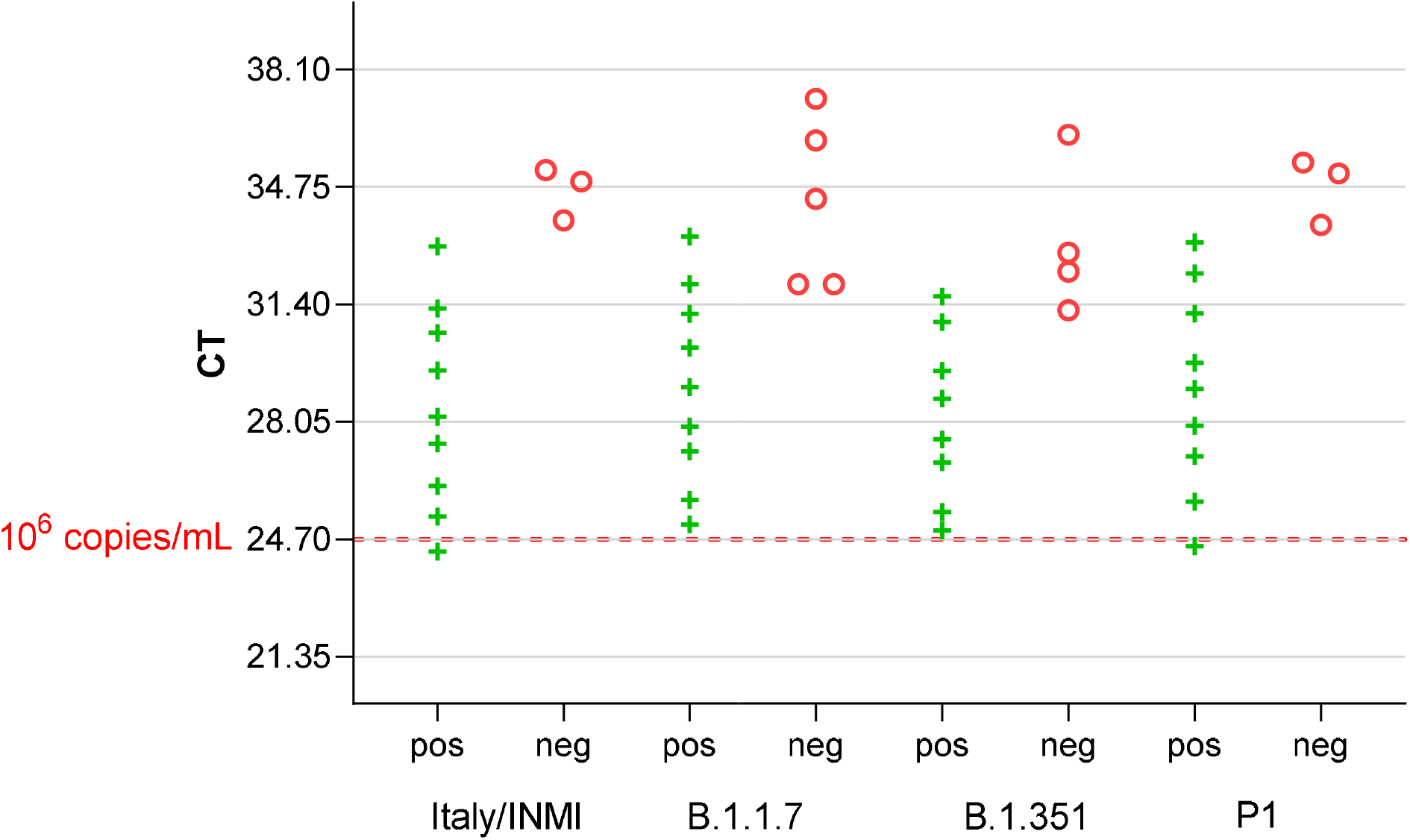
Detectability of SARS-CoV-2 Variants of Concern with the ID NOW COVID-19 system. All analyzed VOC can be detected up to CT values that represent genome loads below 800 per mL.

## Discussion

The ID NOW system is a user-friendly device that does not require laboratory skills for an analysis. It is small and can be regarded as a mobile test device, even if it is not a classical point of care test based on lateral flow assays. The total run time for a negative result is approximately 13 minutes and less than 10 minutes for a positive result. The sample throughput is low as each device can only process one sample at a time. Apart from this, it shows several advantages compared to common point of care tests. The ID NOW system has an analytical sensitivity which is close to that of real-time PCR systems. Compared to the reference real-time PCR assay used in this study, which can detect down to <200 genome copies per mL specimen, the ID NOW has a 50% detection probability of 3,700 genome copies per mL and can detect down to 800 genome copies in individual specimens. The specificity of the ID NOW system was 100% when SARS-CoV-2-negative specimens were analyzed, as well as specimens containing respiratory viruses including influenza viruses and coronaviruses.

Considering the turn-around time of a real-time PCR diagnostic test, the ID NOW has significant benefits when low sample numbers have to be analyzed. Therefore, all settings in which rapid results for few specimens are required, the ID NOW provides reliable and sensitive results. For example, considering that there are medical settings in which a patient cannot wear a face-mask, e.g. at the dentist or the otorhinolaryngologist, the process of excluding infectivity for this patient would take less than 15 minutes. In addition, the ID NOW offers also the testing for influenza A and B, RSV and group A streptococci. Depending on what is required, with the option to use swabs directly as well as liquid (generated by shaking the swab in buffer), one specimen of a patient could be analyzed for additional pathogens, either in parallel by using multiple devices or, considering longer time to result, step by step one after the other. Summing up, the ID NOW COVID-19 test is a useful piece in the puzzle of diagnostic tools that are applied to identify SARS-CoV-2 infections.

The authors are grateful to the diagnostic team at RKI for analyzing the specimens by routine real-time PCR and to Ursula Erikli for copy-editing.

## Supporting information

Highlights

## Data Availability

Data have been collected in Excel format.

## Authors contributions

Eva Krause, Andreas Puyskens and Janine Michel prepared and analyzed the specimens as well as the data. Daniel Bourquain provided variants of concern and analyzed the data. Annika Brinkmann sequenced the VOC genomes. Barbara Biere provided and quantified the specificity panel. Lars Schaade conceptualized the study. Andreas Nitsche designed the study, analyzed the data and wrote the manuscript

## References

[1] WHO/WHE/2021.02, COVID-19 Strategic Preparedness and Response Plan (SPRP 2021), 2021.

[2] Y.W. Tang, J.E. Schmitz, D.H. Persing, C.W. Stratton, Laboratory diagnosis of COVID-19: Current issues and challenges, J. Clin. Microbiol. 58 (2020). https://doi.org/10.1128/JCM.00512-20.

[3] I.M. Mackay, K.E. Arden, A. Nitsche, Real-time PCR in virology., Nucleic Acids Res. 30 (2002) 1292–1305. https://doi.org/10.1093/nar/30.6.1292.

[4] D.K.W. Chu, Y. Pan, S.M.S. Cheng, K.P.Y. Hui, P. Krishnan, Y. Liu, D.Y.M. Ng, C.K.C. Wan, P. Yang, Q. Wang, M. Peiris, L.L.M. Poon, Molecular Diagnosis of a Novel Coronavirus (2019-nCoV) Causing an Outbreak of Pneumonia., Clin. Chem. 66 (2020) 549–555. https://doi.org/10.1093/clinchem/hvaa029.

[5] V.M. Corman, O. Landt, M. Kaiser, R. Molenkamp, A. Meijer, D.K.W. Chu, T. Bleicker, S. Brünink, J. Schneider, M.L. Schmidt, D.G.J.C. Mulders, B.L. Haagmans, B. Van Der Veer, S. Van Den Brink, L. Wijsman, G. Goderski, J.L. Romette, J. Ellis, M. Zambon, M. Peiris, H. Goossens, C. Reusken, M.P.G. Koopmans, C. Drosten, Detection of 2019 novel coronavirus (2019-nCoV) by real-time RT-PCR, Eurosurveillance. (2020). https://doi.org/10.2807/1560-7917.ES.2020.25.3.2000045.

[6] M.J. Loeffelholz, Y.W. Tang, Laboratory diagnosis of emerging human coronavirus infections–the state of the art, Emerg. Microbes Infect. 9 (2020) 747–756. https://doi.org/10.1080/22221751.2020.1745095.

[7] M.L. Holshue, C. DeBolt, S. Lindquist, K.H. Lofy, J. Wiesman, H. Bruce, C. Spitters, K. Ericson, S. Wilkerson, A. Tural, G. Diaz, A. Cohn, L. Fox, A. Patel, S.I. Gerber, L. Kim, S. Tong, X. Lu, S. Lindstrom, M.A. Pallansch, W.C. Weldon, H.M. Biggs, T.M. Uyeki, S.K. Pillai, First Case of 2019 Novel Coronavirus in the United States, N. Engl. J. Med. 382 (2020) 929–936. https://doi.org/10.1056/nejmoa2001191.

[8] J.F.W. Chan, S. Yuan, K.H. Kok, K.K.W. To, H. Chu, J. Yang, F. Xing, J. Liu, C.C.Y. Yip, R.W.S. Poon, H.W. Tsoi, S.K.F. Lo, K.H. Chan, V.K.M. Poon, W.M. Chan, J.D. Ip, J.P. Cai, V.C.C. Cheng, H. Chen, C.K.M. Hui, K.Y. Yuen, A familial cluster of pneumonia associated with the 2019 novel coronavirus indicating person-to-person transmission: a study of a family cluster, Lancet. 395 (2020) 514–523. https://doi.org/10.1016/S0140-6736(20)30154-9.

[9] FIND evaluation update: SARS-CoV-2 molecular diagnostics - FIND, (2021).

[10] J. Dinnes, J.J. Deeks, S. Berhane, M. Taylor, A. Adriano, C. Davenport, S. Dittrich, D. Emperador, Y. Takwoingi, J. Cunningham, S. Beese, J. Domen, J. Dretzke, L. Ferrante di Ruffano, I.M. Harris, M.J. Price, S. Taylor-Phillips, L. Hooft, M.M. Leeflang, M.D. McInnes, R. Spijker, A. Van den Bruel, Rapid, point-of-care antigen and molecular-based tests for diagnosis of SARS-CoV-2 infection, Cochrane Database Syst. Rev. (2021). https://doi.org/10.1002/14651858.CD013705.pub2.

[11] FIND evaluation of SARS-CoV-2 antigen (Ag) detecting tests - FIND, (n.d.).

[12] Paul-Ehrlich-Institut - Coronavirus und COVID-19 - SARS-CoV-2-Testsysteme, (n.d.).

[13] K.M. Koczula, A. Gallotta, Lateral flow assays, Essays Biochem. 60 (2016) 111–120. https://doi.org/10.1042/EBC20150012.

[14] Abbott Inc., Abbott ID NOWTM COVID-19, (2021).

[15] A. Fritsch, B. Schweiger, B. Biere, Influenza c virus in pre-school children with respiratory infections: Retrospective analysis of data from the national influenza surveillance system in germany, 2012 to 2014, Eurosurveillance. 24 (2019). https://doi.org/10.2807/1560-7917.ES.2019.24.10.1800174.

[16] R. Wölfel, V.M. Corman, W. Guggemos, M. Seilmaier, S. Zange, M.A. Müller, D. Niemeyer, T.C. Jones, P. Vollmar, C. Rothe, M. Hoelscher, T. Bleicker, S. Brünink, J. Schneider, R. Ehmann, K. Zwirglmaier, C. Drosten, C. Wendtner, Virological assessment of hospitalized patients with COVID-2019, Nature. 581 (2020) 465–469. https://doi.org/10.1038/s41586-020-2196-x.

[17] J. Bullard, K. Dust, D. Funk, J.E. Strong, D. Alexander, L. Garnett, C. Boodman, A. Bello, A. Hedley, Z. Schiffman, K. Doan, N. Bastien, Y. Li, P.G. van Caeseele, G. Poliquin, Predicting infectious severe acute respiratory syndrome coronavirus 2 from diagnostic samples, Clin. Infect. Dis. 71 (2020) 2663–2666. https://doi.org/10.1093/cid/ciaa638.

[18] J.J.A. van Kampen, D.A.M.C. van de Vijver, P.L.A. Fraaij, B.L. Haagmans, M.M. Lamers, N. Okba, J.P.C. van den Akker, H. Endeman, D.A.M.P.J. Gommers, J.J. Cornelissen, R.A.S. Hoek, M.M. van der Eerden, D.A. Hesselink, H.J. Metselaar, A. Verbon, J.E.M. de Steenwinkel, G.I. Aron, E.C.M. van Gorp, S. van Boheemen, J.C. Voermans, C.A.B. Boucher, R. Molenkamp, M.P.G. Koopmans, C. Geurtsvankessel,A.A. van der Eijk, Duration and key determinants of infectious virus shedding in hospitalized patients with coronavirus disease-2019 (COVID-19), Nat. Commun. 12 (2021). https://doi.org/10.1038/s41467-020-20568-4.

