## Supplementary material for "Sensitive on-site detection of SARS-CoV-2 by ID NOW COVID-19": Highlights

The ID NOW COVID-19 device enables SARS-CoV-2 genome detection down to 800 genome copies per mL, and the 50% detection probability is 3,700 genome copies per mL.

Time to result is approximately 13 minutes for negative specimens. The analytical specificity is 100% as shown by negative specimens and specimens containing 16 different respiratory viruses.
